# Circular RNA detection identifies *circPSEN1* alterations in brain specific to Autosomal Dominant Alzheimer Disease

**DOI:** 10.1101/2021.10.29.21265617

**Authors:** Hsiang-Han Chen, Abdallah Eteleeb, Ciyang Wang, Maria Victoria Fernandez, John P. Budde, Kristy Bergmann, Joanne Norton, Fengxian Wang, Curtis Ebl, John C. Morris, Richard J. Perrin, Randall J. Bateman, Eric McDade, Chengjie Xiong, Alison Goate, Martin Farlow, Jasmeer Chhatwal, Peter R Schofield, Helena Chui, Oscar Harari, Carlos Cruchaga, Laura Ibanez, Dominantly Inherited Alzheimer Network

## Abstract

**Background:** Autosomal-dominant Alzheimer’s disease (ADAD) is caused by pathogenic mutations in *APP, PSEN1*, and *PSEN2*, which usually lead to an early age at onset (<65). Circular RNAs are a family of non-coding RNAs highly expressed in the nervous system and especially in synapses. We aimed to investigate differences in brain gene expression of linear and circular transcripts from the three ADAD genes in controls, sporadic AD, and ADAD.

**Methods:** We obtained and sequenced RNA from brain cortex using standard protocols. Linear counts were obtained using the TOPMed pipeline; circular counts, using python package DCC. After stringent quality control (QC), we obtained the counts for *PSEN1, PSEN2* and APP genes. Only circ*PSEN1* passed QC. We used DESeq2 to compare the counts across groups, correcting for biological and technical variables. Finally, we performed *in-silico* functional analyses using the Circular RNA interactome website and DIANA mirPath software.

**Results:** Our results show significant differences in gene counts of circ*PSEN1* in ADAD individuals, when compared to sporadic AD and controls (ADAD=22, AD=274, Controls=25 – ADADvsCO: log2FC=0.786, p=9.08×10-05, ADADvsAD: log2FC=0.576, p=2.00×10-03). The high gene counts are contributed by two circ*PSEN1* species (hsa_circ_0008521 and hsa_circ_0003848). No significant differences were observed in linear *PSEN1* gene expression between cases and controls, indicating that this finding is specific to the circular forms. In addition, the high circ*PSEN1* levels do not seem to be specific to *PSEN1* mutation carriers; the counts are also elevated in APP and *PSEN2* mutation carriers. *In-silico* functional analyses suggest that circ*PSEN1* is involved in several pathways such as axon guidance (p=3.39×10^−07^), hippo signaling pathway (p=7.38×10^−07^), lysine degradation (p=2.48×10^−05^) or Wnt signaling pathway (p=5.58×10^−04^) among other KEGG pathways. Additionally, circ*PSEN1* counts were able to discriminate ADAD from sporadic AD and controls with an AUC above 0.70.

**Conclusions:** Our findings show the differential expression of circ*PSEN1* is increased in ADAD. Given the biological function previously ascribed to circular RNAs and the results of our *in-silico* analyses, we hypothesize that this finding might be related to neuroinflammatory events that lead or that are caused by the accumulation of amyloid-beta.

## 1. Background

Alzheimer disease (AD) is the most common cause of dementia; approximately 5.8 million Americans suffered from AD in 2019 and, by 2050, it is projected that 14 million individuals in the United States will be affected by AD [1]. AD is characterized by pathological changes in the brain: accumulation of amyloid-beta plaques (extracellular deposits of amyloid-beta peptides) and neurofibrillary tangles (NFTs, intraneuronal fibrillar aggregates of hyperphosphorylated tau). Clinically, AD is defined by gradual and progressive memory loss [2]. AD can be categorized as sporadic AD or Autosomal Dominant AD (ADAD) [3]. ADAD is caused by mutations or duplications of the amyloid precursor protein (*APP*), mutations in presenilin 1 (*PSEN1*), or mutations in presenilin 2 (*PSEN2*), an autosomal dominant inheritance within family members for more than two generations, and an onset earlier than 65 years old [4, 5]. More than 400 mutations have been reported on these three genes, but *PSEN1* harbors the most mutations, which are also associated with the youngest age at onset with affected individuals typically being 30-50 years old [4, 6-8]. In fact, *PSEN1* mutations have also been reported in late-onset AD [9]. Even though ADAD is rare (<0.5% of all AD cases) [10], these cases have provided unique insights into the pathobiology of the disease, especially the formulation of the amyloid hypothesis: that accumulation of amyloid-beta aggregates initiates the pathologic process of AD. All three ADAD-causing genes are part of the amyloid-beta processing pathway. However, neuropathological studies have shown that there are common and distinct pathological characteristics [11]. Both ADAD and AD present neuronal loss, neurofibrillary tangles, amyloid plaques and cerebral amyloid angiopathy among others, but ADAD show, for example, cottonwool plaques, more severe cerebral amyloid angiopathy, more common intracerebral hemorrhage or higher abundance of Lewy bodies [5, 12].

Recently, studies screening the whole genome or the brain transcriptome have been instrumental in elucidating downstream genes and pathways implicated in disease, highlighting the importance of studying AD beyond the amyloid pathway [13-18]. However, most of these studies have been focused on sporadic AD, so their findings cannot always be extrapolated to Mendelian forms of AD; differences between these two forms of the disease are well known [5, 12, 19]. Several studies focused on ADAD have been limited to genetic studies focused on families [6, 20, 21] and animal studies aiming to understand the amyloid cascade hypothesis [22]. Studies involving large diverse ADAD cohorts are limited.

Circular RNAs are a family of non-coding RNAs that result from backsplicing events (the 3’ end of the transcript links covalently to the 5’ forming a loop) [23, 24]. The knowledge of circular RNAs is still limited, but it is thought that they are implicated in the regulation of microRNAs via sequestration, leading to a loss of function of the microRNA [20, 23-25]. Circular RNAs are highly expressed in the nervous system and especially in synapses [20]. Dysregulation of circular RNAs has already been shown for several central nervous diseases, including AD, Parkinson’s disease, and traumatic brain injury [20, 25-27]. CircRNA were systematically screened in brain samples from AD compared to controls [20]. They successfully identified more than 100 circRNAs associated with AD status and disease severity measured by Braak neurofibrillary tangles (NFT) and Clinical Dementia Rating (CDR®) [28]. When subsetting the analyses to ADAD, 236 circRNAs were found to be dysregulated; 56 of them independently of the severity measured by Braak NFT The circRNAs associated with both ADAD and AD showed larger effect sizes in ADAD than in AD. However, no specific analyses regarding the circular forms of the ADAD genes were performed. In this study, we used bulk RNA-seq to postmortem parietal cortex samples from controls, sporadic AD, and ADAD, we investigated the gene expression profiles of linear and circular transcripts of the ADAD genes to determine their possible involvement in the pathobiology of ADAD.

## 2. Methods

### 2.1 Study Population

The discovery phase included bulk RNA-seq data from parietal cortex samples from non-Hispanic white (NHW) participants: 18 ADAD participants from the Dominantly Inherited Alzheimer Network (DIAN) (14 *PSEN1* and four APP mutation carriers), 59 sporadic AD participants and ten control participants from the Knight-ADRC at Washington University in Saint Louis. The replication phase included four ADAD cases from DIAN (two *PSEN1*, one *PSEN2*, and one APP *mutation* carriers), and 215 sporadic AD cases and 15 controls from the Knight-ADRC (Table 1). Finally, we leveraged the Mount Sinai Brain Bank (MSBB) dataset (syn3157743) for replication of the sporadic AD findings (Supplementary Table 1). MSBB contains brain RNA-seq data from different brain regions. From Brodmann area (BM) 10 (frontal pole): 143 AD, 29 Controls; from BM22 (superior temporal gyrus): 134 AD, 26 Controls; from BM36 (parahippocampal gyrus): 123 AD, 24 Controls; from (BM) 44 (inferior frontal gyrus): 132 AD, 25 Controls.

**Table 1:** Demographic Characteristics of the Discovery and Replication Datasets

|  | Discovery |  |  | Replication |  |  | Joint Dataset |  |  |
| --- | --- | --- | --- | --- | --- | --- | --- | --- | --- |
|  | ADAD | AD | Controls | ADAD | AD | Controls | ADAD | AD | Controls |
| **N** | 18 | 59 | 10 | 4 | 215 | 15 | 22 | 274 | 25 |
| **Age at Death (years)** | 54.0<br>(38.8-88.1) | 84.0<br>(72.0-96.1) | 92.5<br>(75.7-103.4) | 58.0<br>(42.1-75.6) | 83.0<br>(68.0-98.0) | 87.0<br>(80.0-100.5) | 54.0<br>(39.0-87.5) | 83.0<br>(68.7-98.0) | 90.0<br>(79.2-103.0) |
| **Sex (% males)** | 66% | 49% | 30% | 75% | 38% | 40% | 68% | 40% | 36% |
| **Age at Onset (years)** | 39.0<br>(28.6-76.0) | 75.0<br>(58.8-90.2) | - | 62.0<br>(52.1-71.9) | 73.0<br>(55.6-92.0) | - | 51.0<br>(28.9-75.8) | 73.0<br>(56.0-92.0) | - |
| **PSEN1 Mutation (% of carriers)** | 77% | 0% | 0% | 50% | 0% | 0% | 72% | 0% | 0% |
| **PMI (hours)** | 9.2<br>(4.2-29.7) | 11.6<br>(4.4-22.5) | 10.5<br>(5.1-19.4) | 10.3<br>(4.7-14.9) | 12.0<br>(4.0-23.0) | 9.1<br>(3.0-21.0) | 9.2<br>(4.5-26.2) | 12.0<br>(4.0-22.7) | 9.5<br>(3.2-21.0) |
| **CDR® at Death** | 2.0<br>(0.7-3.0) | 3.0<br>(1.0-3.0) | 0.3<br>(0-0.8) | 3.0<br>(2.1-3.0) | 3.0<br>(0.5-3.0) | 0<br>(0-0.5) | 2.5<br>(0.7-3.0) | 3.0<br>(0.5-3.0) | 0<br>(0-0.5) |
| **Braak NFT** | 6.0<br>(5.0-6.0) | 5.5<br>(2.6-6.0) | 2.0<br>(1.0-3.2) | 6.0<br>(6.0-6.0) | 6.0<br>(2.0-6.0) | 2.0<br>(1.4-3.0) | 6.0<br>(5.0-6.0) | 6.0<br>(2.0-6.0) | 2.0<br>(1.0-3.0) |

ADAD=Autosomal Dominant Alzheimer Disease; AD=Sporadic Alzheimer Disease; PSEN1= Presenilin1; PMI=Post-Mortem Interval; CDR®=Clinical Dementia Rating; NFT=NeuroFilament Tangles. Age at Death and Age at Onset (AAO) are expressed as median age (95% Inter Quartile Interval).

### 2.2 Library Preparation and Sequencing

The data generation for the discovery dataset has been previously published [20, 21]. We followed the same protocol to generate the replication dataset. Briefly, total RNA was obtained from frozen parietal cortex tissue using the Tissue Lyser LT and RNeasy Mini Kit (Qiagen). After quality control, libraries were generated using TruSeq Stranded Total RNA Sample Prep with Ribo-Zero Gold kit (Illumina). Eighty million 2×150bp reads were generated on average for each sample using an Illumina HiSeq 4000. For the replication dataset, we obtained the RNA from frozen brain tissue with the Maxwell RSC simplyRNA tissue kit (Promega). After quality control, TruSeq Stranded Total RNA Sample Prep with Ribo-Zero Gold kit (Illumina) was used to generate the libraries. Thirty-five million 2×150bp reads were generated on average for each sample using an Illumina NovaSeq 6000.

### 2.3 RNA-seq Quality Control, Alignment, and Circular RNAs detection

Both datasets were processed and aligned separately following similar pipelines as the ones previously published by our group [20, 21]. Genome reference and gene models were selected following the TOPMed pipeline (https://github.com/broadinstitute/gtex-pipeline/blob/master/TOPMed_RNAseq_pipeline.md). Reference genome GRCh38 and GENCODE 33 annotation, including the addition of ERCC spike-in annotations were used. We excluded ALT, HLA, and Decoy contigs from the reference genome due to the lack of RNA-seq tools that allow proper handling of these regions. To obtain the linear counts we followed standard guidelines. Briefly, the raw reads were aligned to the human reference genome (GRCh38) using STAR (v.2.7.1a) [29]. We evaluated the quality of the alignment using sequencing metrics such as reads distribution, ribosomal content or alignment quality provided by STAR [29] using Picard tools (v.2.8.2) [30]. Gene expression was quantified using Salmon (v.1.2.0) [31] and the GENCODE reference genome (GRCh38). All transcripts or genes with less than ten reads in more than 90% of the individuals were removed.

To obtain the circular RNA counts, all raw reads were first aligned to the human reference genome (GRCh38) using STAR [29] in chimeric alignment mode. The remaining alignment parameters were selected specifically for circular RNA detection as suggested by the developers of the circular RNA calling software DCC [32]. Circular RNA detection, annotation and quantification was performed using DCC (v.0.4.8). We excluded any circRNA that had missing counts in more than 25% of the samples. As part of the general quality control, circRNAs that were not present in at least three samples, with a minimum of three counts in at least one of them, were removed. Additionally, we removed any circRNA with missing counts in more than 75% of the samples. Then we proceeded to extract the linear and circular forms corresponding to the three ADAD genes (APP, *PSEN1* and *PSEN2*) from each dataset. For all three datasets, only the circ*PSEN1* could be detected; circ*APP* and circ*PSEN2* were not detected. As a consequence, the analyses were focused on linear *PSEN1* and circ*PSEN1*. The species of circ*PSEN1* varied depending on the dataset and the brain region for the MSBB. Overall, seven circ*PSEN1* species were detected by DCC; but four of them (hsa_circ_0008521, hsa_circ_0003848, hsa_circ_0007013, hsa_circ_0002564) were commonly detected in all three datasets and most of the brain areas (except hsa_circ_0008521 for BM10, hsa_circ_0007013 for BM22 and BM44).

### 2.4 Statistical Analyses

We tested if the levels of circular *PSEN1* (circ*PSEN1*) and linear *PSEN1* were different among groups by comparing AD and control participants, ADAD and AD participants, and ADAD and control participants in both the discovery and the replication datasets. The same analysis of circ*PSEN1* was also performed between sporadic AD participants and control subjects in the MSBB dataset (no ADAD participants were available in the MSBB dataset). We also investigated which specific circ*PSEN1* transcripts were predominant in each group and dataset. After normalization of the counts, differential expression (DE) analyses were performed specifically for circ*PSEN1* and linear *PSEN1* using DEseq2 version 1.22.2 [33] to determine significance. Any association with p-value<0.05 was considered significant. All DE analyses were adjusted for postmortem interval (PMI), RNA quality as measured by median transcript integrity number (TIN) [34] and sex. We also tested if circ*PSEN1* was associated to Braak NFT or age at death to investigate if our findings were driven by disease severity as previously descrived [20]. Briefly, the variable of interest was added to the model to evaluate the effect on the p-value, effect size and direction of the circ*PSEN1* association.

Due to the moderate size of our sample, we combined the discovery and replication datasets, since they were processed using the same pipeline, and performed a joint analysis adding dataset to the model to adjust for possible differences in the DE analysis.

### 2.5. *In-Silico* Functional Study

To investigate the biological function of circ*PSEN1*, we accessed the Circular RNA Interactome website [35] to predict which miRNAs have the potential to target any of the circ*PSEN1* species identified in our datasets. Then, we used the DIANA mirPath software version 3 [36] to identify which genes and pathways were regulated by the identified miRNA using the microT-CDS algorithm and the Kyoto Encyclopedia of Genes and Genomes (KEGG). Finally, we investigated if any of the common genes listed in the pathways identified by the DIANA mirPath software version 3 were differentially expressed in the ADAD cases compared to controls or sporadic AD cases in the discovery dataset.

### 2.6. Discriminative Ability of circ*PSEN1*

To evaluate if circ*PSEN1* can discriminate ADAD from the other brains, we used three binomial regression models built using three different circ*PSEN1* normalized counts: aggregate counts, hsa_circ_0008521, and hsa_circ_0003848 counts to classify ADAD vs. controls, ADAD vs. AD, and AD vs. controls in both the discovery and the replication datasets separately. We used the discovery dataset to train the models, and the replication dataset to validate them. We then evaluated the model performance through receiver operating characteristic curves (ROCs) and areas under the ROC curve (AUCs). The binomial regression models and the receiver operating characteristic curve analyses were performed using R packages stats version 3.5.2 and ROCR version 1.0-7.

## 3. Results

### 3.1 circ*PSEN1* is more abundant in ADAD than Sporadic AD and Controls

The circ*PSEN1* normalized counts were significantly higher in ADAD cases (N=18) compared to controls (N=10; p=1.61×10^−04^, log_2_FC=0.81) and sporadic AD cases (N=59; p=5.00×10^−03^, log_2_FC=0.52), but did not differ between AD cases and controls (p=0.21, log_2_FC=0.29) (Table 2 and Figure 1A) in the discovery dataset. The trend was also observed in the replication dataset in all three comparisons (log_2_FC_ADADvsCO_=0.65; log_2_FC_ADADvsAD_=0.64; log_2_FC_ADvsCO_=0.10) (Table 2 and Figure 1B). Due to the limited sample size of the ADAD group, we do not have enough statistical power to detect differences in the replication dataset (24 ADAD samples are required to have 80% power with a probability of type I error of 0.05). Consequently, we performed a joint analysis of the two datasets. A more significant association was found in the join analyses than in the discovery dataset, indicating that the higher expression level of circ*PSEN1* specific to ADAD than sporadic AD cases and controls (p_ADADvsCO_ =9.08×10^−05^, log_2_FC=0.79; p_ADADvsAD_ =2.00×10^−03^, log_2_FC=0.58; p_ADADvsCO_ =0.44, log_2_FC=0.14; Table 2).

**Table 2:** Differential Expression Results

|  |  | **Circular *PSEN1*** |  | **Linear *PSEN1*** |  |
| --- | --- | --- | --- | --- | --- |
|  |  | log<sub>2</sub>FC | P Value | log<sub>2</sub>FC | P Value |
| **ADAD vs Controls** | Discovery | 0.811 | **1.61×10<sup>-04</sup>** | 0.192 | 0.107 |
|  | Replication | 0.653 | 0.168 | -0.191 | 0.442 |
|  | Joint Analysis | 0.786 | **9.08×10<sup>-05</sup>** | 0.080 | 0.489 |
| **ADAD vs AD** | Discovery | 0.523 | **5.00×10<sup>-03</sup>** | 0.144 | 0.172 |
|  | Replication | 0.644 | 0.198 | -0.139 | 0.583 |
|  | Joint Analysis | 0.576 | **2.00×10<sup>-03</sup>** | 0.064 | 0.568 |
| **AD vs Controls** | Discovery | 0.289 | 0.208 | 0.101 | 0.434 |
|  | Replication | 0.102 | 0.708 | -0.153 | 0.247 |
|  | Joint Analysis | 0.142 | 0.441 | -0.061 | 0.540 |

ADAD=Autosomal Dominant Alzheimer Disease; AD=Sporadic Alzheimer Disease; *PSEN1*=Presenilin 1.

**Figure 1.**
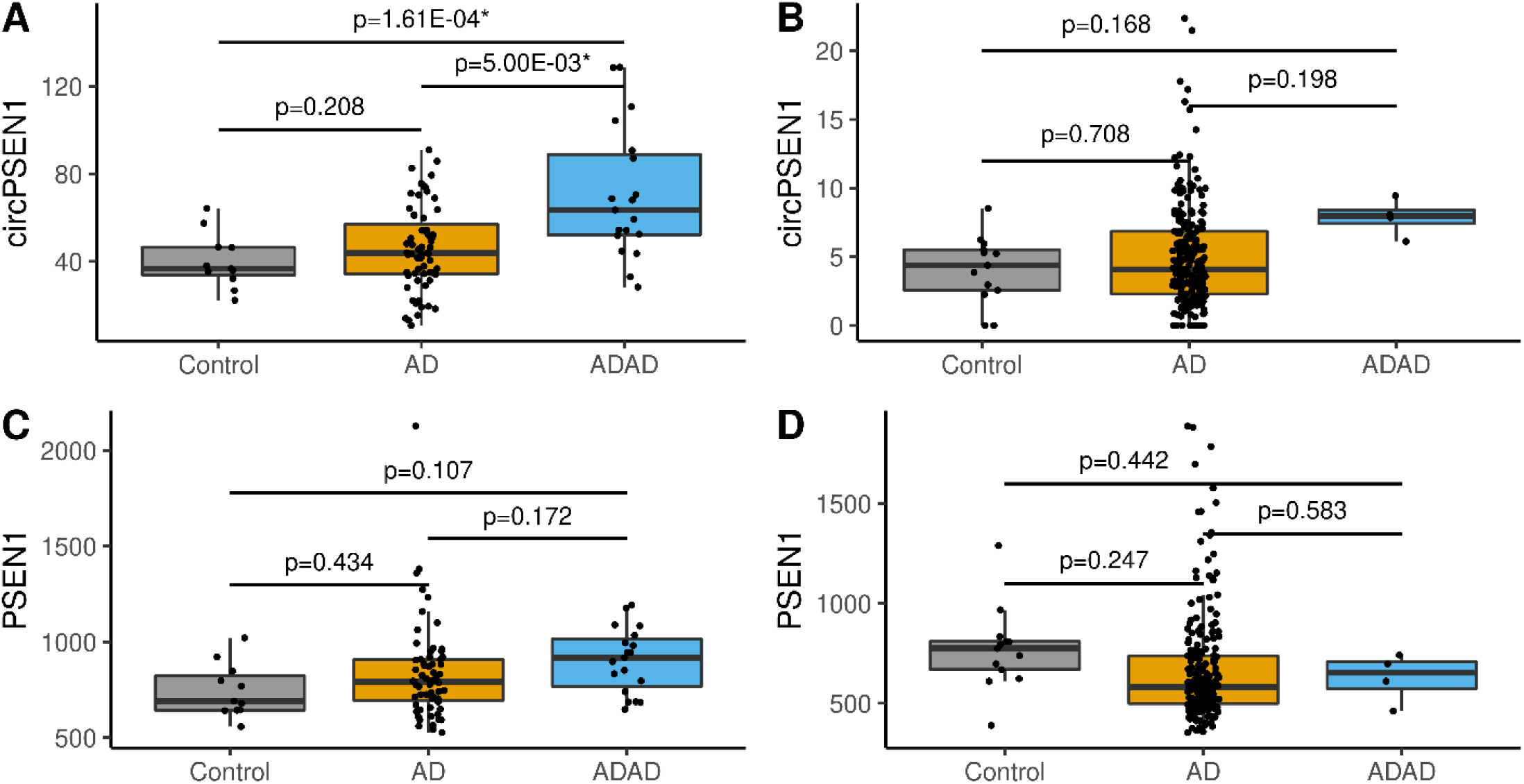
Comparison of the circular *PSEN1* normalized counts in the discovery (Panel A) and replication (Panel B) datasets and the normalized counts for the linear forms of *PSEN1* in the discovery (Panel C) and replication (Panel D) datasets for Controls (grey), AD (Alzheimer disease - ocher) and ADAD (autosomal dominant Alzheimer disease - blue).

We leveraged the MSBB dataset to confirm that there were no differences between the normalized counts of circ*PSEN1* between AD cases and controls. circ*PSEN1* is not differentially expressed in any of the four brain regions (BM10, BM22, BM36, BM44) available in the MSBB dataset (Supplementary Figure 1 and supplementary Table2), which is consistent with our finding that higher circ*PSEN1* expression is specific to ADAD cases.

Similar to RNA-seq, circ*PSEN1* total count is the result of the addition of all the counts from the different species of circ*PSEN1*. We investigated if the differences observed between ADAD and AD or controls were the result of an overall increase or the increase of a specific species. Seven circ*PSEN1* species were found in the discovery and replication dataset (Table 3). All of them are exon derived except one that is intron-exon derived (*circPSEN1*-73147795-73165413). Considering the four species commonly detected in both datasets, the most abundant are hsa_circ_0008521, hsa_circ_0003848, and hsa_circ_0002564; being the first two species significantly different between ADAD and AD (p=6.56×10^−03^, 0.041, log_2_ FC=0.69, 0.54, for joint dataset) and ADAD and Controls (p=1.50×10^−04^, 7.00×10^−03^, log_2_ FC=1.02, 0.77, for joint dataset). Hsa_circ_0002564 was only significantly different between ADAD and AD (p=0.01, log_2_FC=0.60, for joint dataset). No significant differences were found between disease status for hsa_circ_0007013 (Table 3 and Supplementary Figure 2), suggesting that hsa_circ_008521 and hsa_circ_0003848 are driving the association. No significant differences were found between AD and controls for the two species detected in the MSBB dataset in any brain region (Supplementary Table 2). We also investigated the relationship among these three species using correlation tests. We observed high correlation between hsa_circ_0008521 and hsa_circ_0003848 in the discovery (r^2^=0.77, p=2.20×10^−16^), replication (r^2^=0.39, p=3.56×10^−12^), and MSBB (BM22 - r^2^=0.42, p=3.44×10^−08^ | BM36-r^2^=0.58, p=7.79×10^−15^ | BM44-r^2^=0.71, p=2.2×10^−16^) datasets. Hsa_circ_0002564 shows weak correlations with the other two circ*PSEN1* species even though some of them are nominally significant (with hsa_circ_0008521 only in the discovery dataset —r^2^=0.24, p=0.03, and with hsa_circ_0003848 in the discovery—r^2^=0.26, p=0.01 and in the BM22 of the MSBB dataset—r^2^=0.31, p=8.15×10^−05^). This results suggest that hsa_circ_0008521 and hsa_circ_0003848 seem to share the same mechanism of dysregulation, which might be different from hsa_circ_0002564.

**Table 3:** Species of *circPSEN1* characterization and differences across groups

| *circPSEN1* species |  | Discovery |  |  | Replication |  |  | Joint |  |  |
| --- | --- | --- | --- | --- | --- | --- | --- | --- | --- | --- |
|  |  | ADADvsCO | ADADvsAD | ADvsCO | ADADvsCO | ADADvsAD | ADvsCO | ADADvsCO | ADADvsAD | ADvsCO |
| **hsa_circ_0008521** | P value | **5.316×10<sup>-04</sup>** | **0.004** | 0.278 | 0.379 | 0.491 | 0.449 | **1.494×10<sup>-04</sup>** | **6.557×10<sup>-03</sup>** | 0.223 |
|  | log<sub>2</sub> FC | 0.990 | 0.614 | 0.300 | 1.052 | 0.673 | 0.428 | 1.017 | 0.694 | 0.347 |
| **hsa_circ_0003848** | P value | **0.022** | **0.029** | 0.180 | 0.360 | 0.297 | 0.666 | **6.960×10<sup>-03</sup>** | **0.041** | 0.773 |
|  | log<sub>2</sub> FC | 0.701 | 0.436 | 0.295 | 0.972 | 1.148 | -0.266 | 0.769 | 0.536 | 0.082 |
| **hsa_circ_0007013** | P value | 0.796 | 0.795 | 0.386 | 0.811 | 0.750 | 0.780 | 0.843 | 0.704 | 0.499 |
|  | log<sub>2</sub> FC | -0.338 | -0.174 | 0.766 | 0.884 | 0.614 | 0.292 | 0.206 | 0.262 | 0.443 |
| ***circPSEN1***<br>73147795-73165413 | P value | - | - | - | 0.832 | 0.985 | 0.937 | - | - | - |
|  | log<sub>2</sub> FC | - | - | - | -0.833 | -0.186 | -0.375 | - | - | - |
| **hsa_circ_0002564** | P value | 0.066 | **4.336×10<sup>-04</sup>** | 0.706 | 0.636 | 0.656 | 0.769 | 0.059 | **0.012** | 0.585 |
|  | log<sub>2</sub> FC | 0.723 | 0.859 | 0.126 | 0.325 | 0.264 | 0.092 | 0.640 | 0.600 | 0.128 |
| **hsa_circ_0008218** | P value | - | - | - | 0.995 | 0.687 | 0.912 | - | - | - |
|  | log<sub>2</sub> FC | - | - | - | 0.019 | 0.700 | -0.106 | - | - | - |
| **hsa_circ_0032509** | P value | 0.825 | 0.841 | 0.962 | - | - | - | - | - | - |
|  | log<sub>2</sub> FC | 0.700 | 0.624 | 0.207 | - | - | - | - | - | - |

ADAD=Autosomal Dominant Alzheimer Disease; AD=Sporadic Alzheimer Disease; CO=Control; *PSEN1*=Presenilin 1

To ensure that the association was not driven by disease severity or age at death, we tested if Braak NFT score or age at death were influencing the association of circ*PSEN1* with ADAD. We observed no significant changes on the results (Supplementary Table 3), suggesting that these findings are not due to pathology severity or the age of the individual.

### 3.2 Circular *PSEN1* is independent from linear *PSEN1*

We then investigated if the association between ADAD and circ*PSEN1* was also observed in the linear form of *PSEN1*. No significant differences were found (Table 2, Figures 1C and 1D).

We then tested the independency of the normalized counts for the linear and circular forms of *PSEN1*. significant correlation was observed for the ADAD individuals or the controls (Supplementary Figure 3). The correlation between the linear *PSEN1* and the circ*PSEN1* was weak (R_Discovery_=0.40; R_Replication_=0.23), even though it was nominally significant (p_Discovery_ =1.04×10^−03^, p_Replication_ =1.04×10^−03^) in the AD group. To assess if this correlation was affecting our results in the AD group, linear *PSEN1* was added to the differential expression analysis as previously described [37]. The changes of circ*PSEN1* were still significant (p=6.88×10^−04^), even when adjusting for linear *PSEN1*. This result suggests, that even though the correlation of linear and circular *PSEN1* was nominally significant, the linear and circular forms are independent. Similar results were observed for correlation analyses using for *PSEN1* species against linear counts (data not shown).

### 3.3 ADAD individuals have higher circ*PSEN1* counts independently of the mutation

ADAD mutations are most prevalent in *PSEN1* among individuals in both the discovery and the replication datasets. Thus, we evaluated if the high expression levels of circ*PSEN1* were unique to *PSEN1* mutation carriers. In the joint dataset (Figure 2A), the normalized counts of circ*PSEN1* between *PSEN1* mutation carriers (N=16) and APP mutation carriers (N=5) are not significantly different (log2FC=0.245, p=0.414). When compared to AD cases (N=274) or controls (N=25), *PSEN1* mutation carriers showed increased levels of circ*PSEN1* (log_2_FC=0.604, p=0.005, and log_2_FC=0.778, p=8.299×10-04). APP mutation carriers also showed higher counts of circ*PSEN1* when compared to controls (log_2_FC=0.658, p=0.012). Due to the limited sample size of *PSEN2* mutation carriers (N=1), no statistical test was performed. No differences were found among mutations carriers regarding linear *PSEN1* normalized counts (Figure 2B).

**Figure 2.**
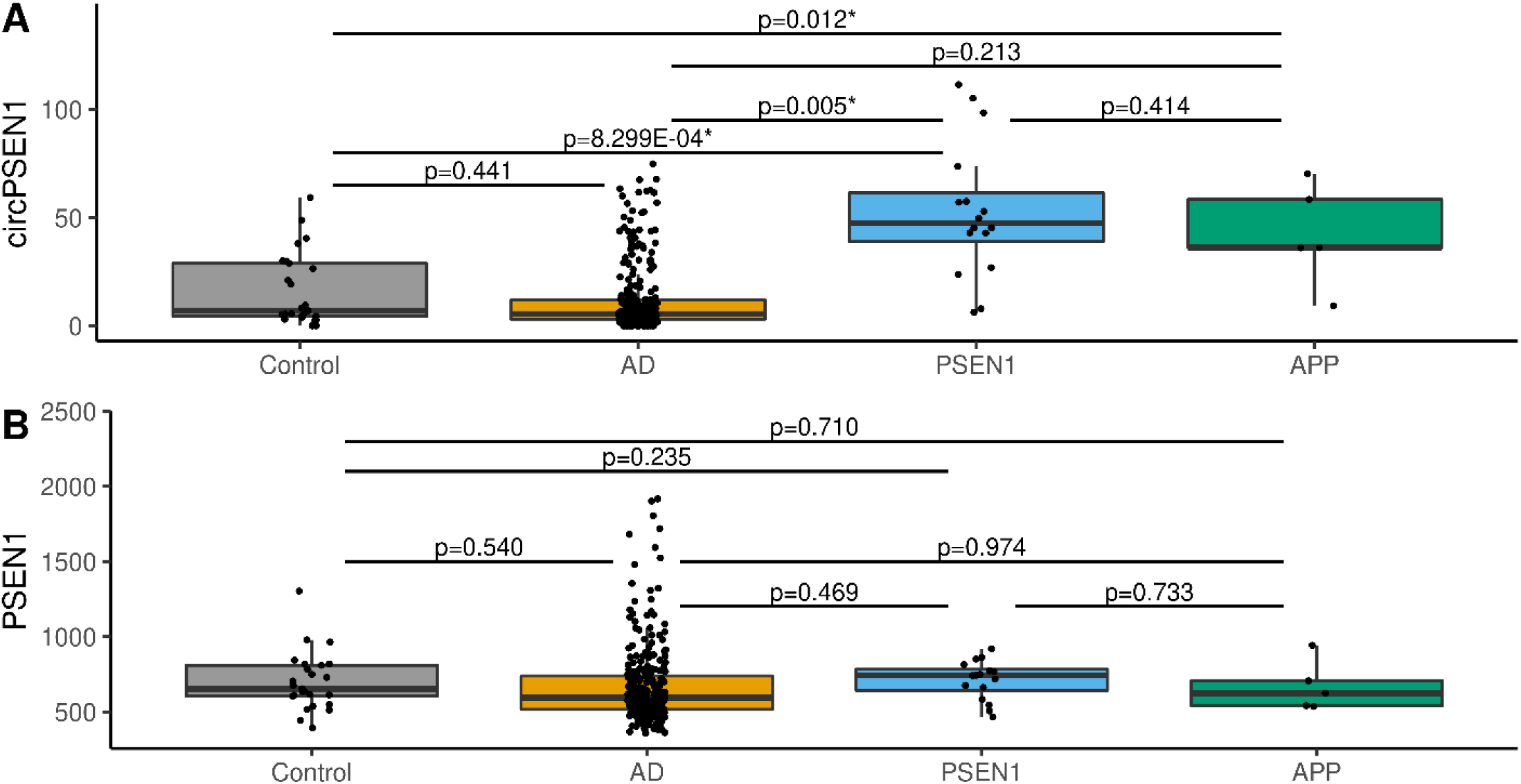
Comparison of the normalized counts of circular *PSEN1*(Panel A)/linear *PSEN1* (Panel B) between controls, AD, and different mutation carriers in the joint dataset - Controls (grey), ADs (Alzheimer disease - ocher), *PSEN1* mutation carriers (blue), and *APP* mutation carriers (green).

### 3.4 *In-silico* analyses functional annotation

We found the three most abundant species of circ*PSEN1* in our samples to be hsa_circ_0008521, hsa_circ_0003848, and hsa_circ_0002564, with the first two most likely driving the overall signal. CircRNAs have been reported to regulate gene expression by sequestering miRNA. Because the biological function of these circ*PSEN1* species has not been described, we explored if the biological function of circ*PSEN1* might be elucidated by those miRNAs targeted to them. Using the Targetscan prediction tool[38] from the CircInteractome database [35], we identified 26 miRNAs that could potentially target to the three most abundant species of circ*PSEN1*. We used them as input in the microT-CDS tool of the DIANA mirPath v.3 software to elucidate which pathways are potentially regulated by the identified miRNA. Several pathways were found significantly associated with these 26 miRNAs: axon guidance (p=3.39×10^−07^); hippo signaling pathway (p=7.38×10^−07^); lysine degradation (p=2.48×10^−05^); and Wnt signaling pathway (p=5.58×10^−04^) among other KEGG pathways (Supplementary Table 4). We identified 31 genes that were common in the top ten KEGG pathways (Supplementary Table 5); the counts of two of them were found significantly lower in ADAD brains compared to AD brains. FDZ4 and RAF1 were nominally significant in the discovery (p=0.037; p=5.55×10^−04^) and replication (p=0.001; p=0.039) datasets. Both genes are related to the transmission of chemical signals between the cell surface and the nucleus.

### 3.5 Circular *PSEN1* normalized counts can discriminate ADAD

The predictive ability of circ*PSEN1* counts (aggregate, hsa_circ_0008521, and hsa_circ_0003848) was evaluated using ROCs and AUCs (Supplementary Figure 4). Aggregated and individual normalized counts of circ*PSEN1* show no predictive ability for AD vs. CO. However, circ*PSEN1* showed good predictive power for ADAD vs. AD. The AUC for the aggregated counts in the discovery dataset was 0.73; that in the replication dataset was 0.79. When we evaluated the predictive power of the two circ*PSEN1* species separately, hsa_circ_0008521 seemed to have slightly better predictive power than hsa_circ_0003848. Yet, the aggregate counts of circ*PSEN1* seemed to show a more robust discriminative power, which generated similar AUCs across datasets.

The discriminative power of circ*PSEN1* increased when we attempted to classify ADAD vs. CO, with an AUC of 0.82 in the discovery dataset and an AUC of 0.85 in the replication dataset for the aggregated counts of circ*PSEN1*. The trends were very consistent for both datasets for all three predictions despite the differences in sample size. This result suggests that, even though the most abundant circ*PSEN1* species seem to have more discriminative power, the less abundant ones (hsa_circ_0007013, hsa_circ_0002564, hsa_circ_0008218, hsa_circ_0032509) are also contributing since the discriminative power of the aggregate counts is more stable in the two datasets.

## 4. Discussion

In this study, we provide evidence that the transcriptional signatures differ between ADAD and AD brains. By analyzing the largest dataset to date of ADAD brains, we have found that circ*PSEN1* expression is increased in ADAD brains but not in AD cases or controls, a result which is independent of age and disease severity (as measured by Braak NFT score). Our results show that this increased expression is not specific to *PSEN1* mutation carriers, as similar results were observed in *APP* mutation carriers, or due to linear *PSEN1*, suggesting a biological mechanism specific to the circ*PSEN1*.

A previous study [20], demonstrated that expression changes of circRNAs in pre-symptomatic AD, sporadic AD, and ADAD are different and not always related to severity of the disease. They found more than 100 circRNAs dysregulated in the context of AD, demonstrating the involvement of circRNA in the pathobiology of AD. In fact, they found even more circRNA dysregulated in their comparison of ADAD participants versus controls. In the present study we add evidence to the importance of the dysregulation of circRNAs. Dube et al, identified that circRNAs that were dysregulated in both AD and ADAD, presented with larger effect sizes in ADAD. In here, we found that circ*PSEN1* that is related to the amyloid pathway is uniquely dysregulated in ADAD participants. This emphasizes the importance of studying not only the molecular similarities between AD and ADAD, but also the differences.

It has been demonstrated that circular RNAs are generated through the spliceosome, suggesting that additional to the miRNA regulation through their sponge function, circular RNA generation is one of the mechanisms that regulates the production of linear RNA [39]. On top of that, spliceosomal proteins have been reported to aggregate with tau tangles [40] and to be down-regulated in the presence of amyloid-beta_42_ [41]. The production of circ*PSEN1* could be due to the mutations present in *PSEN1* via spliceosome alterations. Among the *PSEN1* mutations in this study, p.H163R is located at the boundaries of hsa_circ_0002564. This might explain the dysregulation of this particular circular RNA. However, this mutation was present in only four of the ADAD brain samples, suggesting that other regulatory events are taking place in the ADAD individuals unrelated to this mutation. In fact, previous studies have demonstrated that introns regulate the biogenesis of circular RNA. Given that most of the *PSEN1* mutations are located within the exons, it is likely that this might not be the biological explanation.

Our *in-silico* functional analysis predicted 26 miRNAs that could bind the three abundant circ*PSEN1* species. These 26 miRNAs are significantly associated with several pathways, including wnt, hippo and axon guidance pathways that have been previously related to the development of AD and to neuroinflammation [42-45]. Among the 26 identified miRNAs, miR-144 has previously been associated with AD [46]; in fact, miR-144-3p targets *APP*, significantly inhibiting protein expression [47]. The overexpression of miR-433 targets *JAK2* (janus kinase 2) which contributes to the progression of AD by inhibition of amyloid-beta-induced neuronal viability [48]. Additionally, miR-566 [49] and miR-885-5p [50] were also found to be dysregulated in AD. Finally, miR-655 inhibited the inflammatory response of microglia by targeting *TREM2* (triggering receptor expressed on myeloid 2) [51], which is known to affect amyloid and tau pathologies. Our finding adds evidence to the fact that AD is not restricted to neurons but involves several mechanisms including inflammation [52]. Given the involvement of microglia in the inflammatory process, axon guidance [53], and the role of wnt pathway [45], the presence of circ*PSEN1* might be originating from the microglia. Analyses of circ*PSEN1* using publicly available RNAseq data from IPSCs [54] (data not shown) showed no differences between mutation carriers and isogenic corrected cells. Additionally, previous studies have shown that the neuronal proportion in ADAD brains seems to be lower compared to AD [21]. Together, this suggests that the *circPSEN1* association described in here might not have a neuronal origin.

circ*PSEN1* might be a regulatory factor located at the top hierarchical levels of the dysregulation of the amyloid-beta pathway and leading to the neuroinflammatory status. Our results show that circ*PSEN1* is dysregulated in all ADAD cases, independent of the specific mutation. However, due to the limited sample size of *PSEN2* and *APP* carriers, we cannot disregard the possibility that this alteration is unique to *PSEN1* mutation carriers.

Additional analyses to understand the biological function of circ*PSEN1* and its relationship with *PSEN1* are needed, along with the study of circular and linear forms of *PSEN2* and APP in mutation carriers to elucidate the biological consequences of circular RNAs in ADAD in comparison to AD. If further replicated, circ*PSEN1* might be targeted to diminish neuroinflammation in ADAD individuals to delay the onset of the disease or slow down its progression.

This study includes the largest sample of ADAD brains analyzed to date. However, it is still a study with limited sample size, therefore limiting the statistical power of this analysis. Although our findings are novel and possibly biologically relevant, due to the limited knowledge about circular RNAs and their biological function, along with the relationship between linear and circular forms of the same gene, we cannot claim any causal involvement of circ*PSEN1* with ADAD or AD.

## 5. Conclusions

In conclusion, our circ*PSEN1* differential expression analysis has shown significant differences in the expression of circ*PSEN1* that are unique to ADAD, and independent of gene mutation. Due to the biological function previously ascribed to circular RNAs and our *in-silico* analyses, we hypothesize that this finding might be related to neuroinflammatory events that lead or that are caused by the accumulation of amyloid-beta. Future studies aimed at understanding the biological function of circ*PSEN1* might lead to a better understanding of its pathological involvement with ADAD and its potential as drug-target.

## Data Availability

Discovery dataset data corresponding to the participants who enrolled in the Knight-ADRC can be downloaded at the NIAGADS Knight ADRC collection (NG00083). Additional Knight-ADRC participants and DIAN data is available upon request to qualified researchers. Finally, MSBB dataset is publicly available in Synapse (syn3159438).

## 6. Declarations

### Ethics approval and consent to participate

The Institutional Review Boards of Washington University School of Medicine in St. Louis approved the study, and research was carried out in accordance with the approved protocols. Written informed consent was obtained from all participants or their family members.

### Consent for publication

All authors read and approved the final manuscript.

### Author’s contributions

LI and HHC conceived and wrote this article. AE provided and QCed the data. HHC and CW performed the analyses. AE, CW, MVF, JB, KB, FW, CE, JCM, RJP, RJB, EMD, CX, AG, MF, JC, PRS, HC, OH and CC discussed the project, revised the manuscript and provided critical feedback.

## Acknowledgments

We thank all the participants and their families along with the institutions and all the staff who provided brain tissue, without whom this study would not have been possible. This work was supported by access to equipment made possible by the Hope Center for Neurological Disorders, the Neurogenomics and Informatics Center (NGI: https://neurogenomics.wustl.edu/) and the Departments of Neurology and Psychiatry at Washington University School of Medicine.

This manuscript has been reviewed by DIAN Study investigators for scientific content and consistency of data interpretation with previous DIAN Study publications. We acknowledge the altruism of the participants and their families and contributions of the DIAN research and support staff at each of the participating sites for their contributions to this study.

---

**Supplementary Table 1.** Demographic Characteristics of the Mount Sinai Brain Bank (MSBB) in the different brain regions

|  | BM10 |  | BM22 |  | BM36 |  | BM44 |  |
| --- | --- | --- | --- | --- | --- | --- | --- | --- |
|  | AD | Controls | AD | Controls | AD | Controls | AD | Controls |
| **N** | 143 | 29 | 134 | 26 | 123 | 24 | 132 | 25 |
| **Sex**<br>**(% males)** | 32% | 31% | 0.37 | 0.31 | 0.33 | 0.33 | 0.33 | 0.36 |
| **Age at Onset**<br>**(years)** | 87.0<br>(71.1-90.0) | - | 85.0<br>(71.7-90.0) | - | 87.0<br>(71.1-90.0) | - | 87.0<br>(72.0-90.0) | - |
| **PMI**<br>**(hours)** | 5.0<br>(2.0-15.0) | 10.0<br>(4.0-19.6) | 5.0<br>(2.0-15.3) | 8.5<br>(4.3-19.8) | 5.0<br>(2.0-15.0) | 8.0<br>(3.2-19.0) | 5.0<br>(2.0-15.0) | 9.0<br>(4.0-19.8) |

BM= Brodmann; AD=Sporadic Alzheimer Disease; PMI=Post-Mortem Interval; PMI and Age at Onset is expressed as median age (95% Inter Quartile Interval).

**Supplementary Table 2.** Differential Expression of aggregate circPSEN1 and the species identified in the four MSBB brain regions

|  | Aggregate circPSEN1 |  | hsa_circ_0008521 |  | hsa_circ_0003848 |  |
| --- | --- | --- | --- | --- | --- | --- |
|  | log<sub>2</sub>FoldChange | P value | log<sub>2</sub>FoldChange | P value | log<sub>2</sub>FoldChange | P value |
| **BM10** | 0.029 | 0.877 | - | - | -0.292 | 0.397 |
| **BM22** | 0.029 | 0.901 | 0.448 | 0.374 | 0.317 | 0.381 |
| **BM36** | 0.019 | 0.898 | 0.193 | 0.363 | -0.063 | 0.743 |
| **BM44** | 0.215 | 0.143 | 0.434 | 0.061 | 0.548 | 0.194 |

BM= Brodmann Area; PSEN1=Presinilin1

**Supplementary Table 3.** Differential Expression of circ*PSEN1* when adding, Braak NFT, or age at death to the model to test for the independency of linear and circular forms of *PSEN1*

| Model | Sample Size |  | circPSEN1 DE |  |
| --- | --- | --- | --- | --- |
|  | ADAD | AD | log2FoldChange | P value |
| Status+Sex+PMI+TIN+Dataset+BraakNFT | 17 | 212 | -0.005 | 0.918 |
| Status+Sex+PMI+TIN+Dataset+Age at Death | 22 | 274 | -0.011 | 0.051 |

DE=Differential Expression, ADAD=Autosomal Dominant Alzheimer Disease; AD=Sporadi Alzheimer Disease; PMI=Post-Mortem Interval; TIN=Transcript Integrity Number; NFT=NeuroFibillary Tangles

**Supplementary Table 4.** KEGG pathways identified by DIANA mirPath software to be significantly associated with the miRNA identified by the Circular RNA Interactome to bind circ*PSEN1*.

| KEGG pathway | P value | KEGG pathway | P value |
| --- | --- | --- | --- |
| Proteoglycans in cancer | 6.81E-09 | Adrenergic signaling in cardiomyocytes | 4.47E-03 |
| Axon guidance | 3.39E-07 | Thyroid hormone signaling pathway | 4.47E-03 |
| Hippo signaling pathway | 7.38E-07 | Endometrial cancer | 4.47E-03 |
| Signaling pathways regulating pluripotency of stem cells | 8.99E-06 | Transcriptional misregulation in cancer | 4.47E-03 |
| Lysine degradation | 2.48E-05 | Focal adhesion | 5.04E-03 |
| Pathways in cancer | 1.95E-04 | Neurotrophin signaling pathway | 5.26E-03 |
| ErbB signaling pathway | 3.25E-04 | Non-small cell lung cancer | 6.95E-03 |
| Wnt signaling pathway | 5.58E-04 | Oxytocin signaling pathway | 7.00E-03 |
| Dorso-ventral axis formation | 8.02E-04 | AMPK signaling pathway | 7.98E-03 |
| Hepatitis B | 8.09E-04 | GABAergic synapse | 7.98E-03 |
| Prostate cancer | 8.40E-04 | Glutamatergic synapse | 7.98E-03 |
| Glioma | 8.40E-04 | Adherens junction | 9.07E-03 |
| TGF-beta signaling pathway | 9.47E-04 | Nicotine addiction | 0.01 |
| MAPK signaling pathway | 9.55E-04 | Dopaminergic synapse | 0.01 |
| PI3K-Akt signaling pathway | 1.98E-03 | cAMP signaling pathway | 0.02 |
| Colorectal cancer | 3.16E-03 | Long-term potentiation | 0.02 |
| Ras signaling pathway | 3.16E-03 | Renal cell carcinoma | 0.02 |
| FoxO signaling pathway | 3.18E-03 | Regulation of actin cytoskeleton | 0.02 |
| Biotin metabolism | 4.47E-03 | Circadian entrainment | 0.03 |
| Pancreatic cancer | 4.47E-03 | Melanoma | 0.03 |
| Chronic myeloid leukemia | 4.47E-03 | Rap1 signaling pathway | 0.03 |
| mTOR signaling pathway | 4.47E-03 | Retrograde endocannabinoid signaling | 0.04 |
| Amphetamine addiction | 4.47E-03 |  |  |

**Supplementary Table 5.** Common genes within the KEGG pathways identified by DIANA mirPath software and their differential expression in brains of ADAD, AD and, controls for the discovery and the replication datasets

| Genes | Discovery |  |  |  |  |  | Replication |  |  |  |  |  |
| --- | --- | --- | --- | --- | --- | --- | --- | --- | --- | --- | --- | --- |
|  | ADADvsCO |  | ADADvsAD |  | ADvsCO |  | ADADvsCO |  | ADADvsAD |  | ADvsCO |  |
|  | log<sub>2</sub>FC | P value | log<sub>2</sub>FC | P value | log<sub>2</sub>FC | P value | log<sub>2</sub>FC | P value | log<sub>2</sub>FC | P value | log<sub>2</sub>FC | P value |
| AKT2 | -0.0724 | 6.39E-01 | 0.0739 | 4.49E-01 | -0.1434 | 2.37E-01 | -0.1171 | 6.34E-01 | -0.2327 | 1.55E-01 | 0.1320 | 1.37E-01 |
| GRB2 | 0.0107 | 8.95E-01 | 0.0010 | 9.84E-01 | -0.0087 | 8.99E-01 | 0.0394 | 7.29E-01 | 0.1267 | 2.22E-01 | -0.0709 | 1.93E-01 |
| WNT9B | 0.1527 | 4.46E-01 | 0.0137 | 9.21E-01 | 0.1663 | 3.55E-01 | 0.5069 | 4.16E-01 | 0.8533 | 3.92E-02 | -0.3010 | 1.53E-01 |
| PRKCB | 0.2241 | 2.03E-01 | 0.1816 | 2.38E-01 | -0.0038 | 9.84E-01 | -0.2684 | 6.70E-01 | 0.3839 | 2.53E-01 | -0.6003 | 1.50E-03 |
| SMAD3 | -0.3122 | 1.96E-02 | -0.1087 | 1.97E-01 | -0.1160 | 2.78E-01 | 0.0578 | 8.47E-01 | -0.1531 | 3.73E-01 | 0.0037 | 9.70E-01 |
| WNT10B | 0.4645 | 1.89E-02 | 0.5156 | 9.97E-04 | 0.0411 | 8.37E-01 | 0.1361 | 8.74E-01 | 0.4250 | 2.74E-01 | -0.3501 | 1.14E-01 |
| KRAS | 0.2401 | 5.24E-02 | 0.1162 | 2.21E-01 | 0.1113 | 3.93E-01 | -0.0538 | 8.36E-01 | 0.2019 | 3.13E-01 | -0.1617 | 1.38E-01 |
| MAP2K1 | -0.0121 | 9.45E-01 | -0.0070 | 9.55E-01 | -0.1054 | 4.94E-01 | -0.2347 | 5.05E-01 | 0.2921 | 2.91E-01 | -0.2701 | 7.78E-02 |
| FZD4 | -0.2342 | 2.60E-01 | -0.2418 | 3.74E-02 | 0.0248 | 8.35E-01 | -0.5838 | 4.64E-02 | -0.6329 | 1.35E-03 | -0.0140 | 8.95E-01 |
| PIK3R5 | -1.0713 | 1.01E-04 | -0.5582 | 1.15E-02 | -0.6184 | 2.39E-02 | -0.4223 | 2.62E-01 | -0.6961 | 5.72E-02 | 0.0117 | 9.52E-01 |
| MAPK1 | -0.1731 | 3.46E-02 | -0.1258 | 4.98E-02 | -0.0361 | 6.64E-01 | 0.1213 | 6.07E-01 | 0.1652 | 1.98E-01 | -0.0570 | 4.27E-01 |
| WNT6 | -0.7467 | 2.25E-01 | -0.6210 | 1.37E-01 | -0.6385 | 2.73E-01 | -0.0388 | 9.61E-01 | -1.2207 | 1.22E-01 | 1.1848 | 5.60E-03 |
| CTNNB1 | -0.1878 | 2.36E-03 | -0.0837 | 1.49E-01 | -0.0865 | 2.43E-01 | 0.2385 | 1.91E-01 | 0.0818 | 3.74E-01 | 0.1692 | 7.80E-04 |
| FZD5 | -0.8505 | 5.23E-06 | -0.2045 | 2.10E-01 | -0.6462 | 2.37E-03 | -0.1610 | 7.17E-01 | -0.4072 | 2.37E-01 | 0.3236 | 7.57E-02 |
| WNT5A | -0.1663 | 3.67E-01 | 0.0032 | 9.78E-01 | -0.1675 | 2.02E-01 | 0.3245 | 3.59E-01 | -0.0031 | 9.89E-01 | 0.3022 | 7.92E-03 |
| SMAD4 | -0.6016 | 7.62E-07 | -0.2880 | 1.17E-03 | -0.2960 | 7.09E-03 | -0.1224 | 4.42E-01 | -0.1173 | 3.66E-01 | -0.0067 | 9.23E-01 |
| WNT2B | -0.0399 | 7.89E-01 | 0.0313 | 7.18E-01 | 0.0198 | 8.65E-01 | -0.2666 | 2.93E-01 | 0.0594 | 7.71E-01 | -0.2839 | 8.49E-03 |
| PIK3CG | -0.1939 | 4.56E-01 | 0.0374 | 8.37E-01 | 0.0750 | 7.34E-01 | -0.2287 | 3.92E-01 | -0.2002 | 5.37E-01 | -0.0793 | 6.39E-01 |
| WNT16 | 0.8150 | 5.74E-02 | 0.3080 | 3.71E-01 | 0.2511 | 5.15E-01 | -0.0566 | 9.44E-01 | 0.0844 | 8.90E-01 | -0.4285 | 1.82E-01 |
| PIK3CB | 0.2037 | 1.73E-01 | 0.1259 | 2.29E-01 | 0.0468 | 7.21E-01 | -0.1825 | 3.45E-01 | 0.1385 | 4.86E-01 | -0.2253 | 3.27E-02 |
| PRKCA | -0.1297 | 2.74E-01 | -0.2100 | 1.19E-03 | 0.1436 | 3.46E-02 | -0.0207 | 9.37E-01 | -0.0426 | 7.61E-01 | -0.1032 | 1.92E-01 |
| WNT7A | -0.1159 | 6.37E-01 | -0.2102 | 1.87E-01 | -0.0324 | 8.77E-01 | -1.1896 | 1.74E-02 | -0.2755 | 3.60E-01 | -0.9619 | 2.37E-08 |
| GSK3B | 0.1766 | 4.39E-02 | 0.1006 | 2.28E-01 | 0.0691 | 5.16E-01 | -0.0495 | 7.75E-01 | 0.2256 | 1.69E-01 | -0.1830 | 3.72E-02 |
| AKT3 | 0.2262 | 2.32E-02 | 0.0977 | 2.36E-01 | 0.1178 | 2.91E-01 | 0.0368 | 7.22E-01 | 0.2038 | 1.41E-01 | -0.0873 | 2.32E-01 |
| PIK3CA | 0.0970 | 3.85E-01 | 0.0426 | 5.74E-01 | 0.0530 | 6.02E-01 | -0.1083 | 4.30E-01 | 0.1183 | 3.53E-01 | -0.1586 | 1.88E-02 |
| RAF1 | -0.3583 | 1.22E-05 | -0.1794 | 5.55E-04 | -0.2195 | 2.22E-04 | -0.1442 | 2.62E-01 | -0.2037 | 3.87E-02 | 0.0670 | 1.96E-01 |
| WNT9A | -0.1131 | 8.09E-01 | 0.0392 | 8.88E-01 | 0.0057 | 9.86E-01 | -0.2204 | 7.69E-01 | -0.6735 | 1.61E-01 | 0.1772 | 4.97E-01 |
| FZD8 | -0.5680 | 3.83E-02 | -0.3553 | 7.55E-02 | -0.1821 | 4.65E-01 | -0.7128 | 1.31E-01 | -0.5984 | 9.57E-02 | -0.3548 | 6.93E-02 |
| FZD1 | -0.5506 | 1.36E-02 | -0.2030 | 1.72E-01 | -0.3550 | 4.23E-02 | -0.3247 | 4.83E-01 | -0.5988 | 2.16E-02 | 0.1954 | 1.77E-01 |
| FZD3 | 0.1273 | 3.53E-01 | 0.0992 | 3.57E-01 | 0.0435 | 7.50E-01 | -0.0343 | 9.25E-01 | 0.1636 | 4.44E-01 | -0.1864 | 1.10E-01 |
| FZD6 | 0.1862 | 3.27E-01 | 0.1782 | 1.69E-01 | 0.0854 | 6.02E-01 | 0.0788 | 8.32E-01 | -0.3292 | 1.94E-01 | 0.2061 | 1.22E-01 |

**Supplementary Figure 1.**
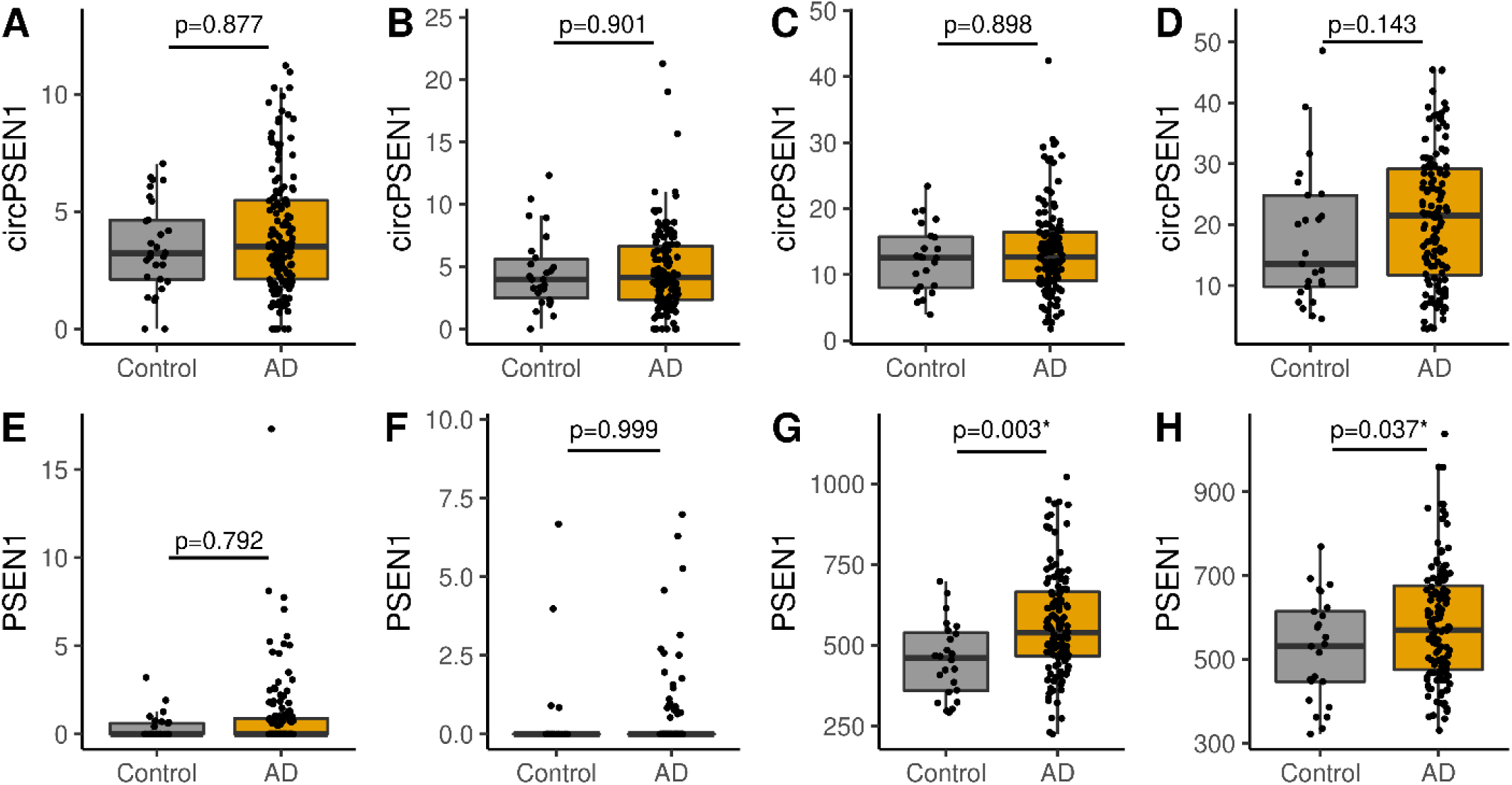
Circular and linear *PSEN1* normalized counts in the Mount Sinai Brain Bank dataset. Panels A to D represent circ*PSEN1* and panels E to H linear *PSEN1* (BM10, BM22, BM36 and BM44 respectively).

**Supplementary Figure 2.**
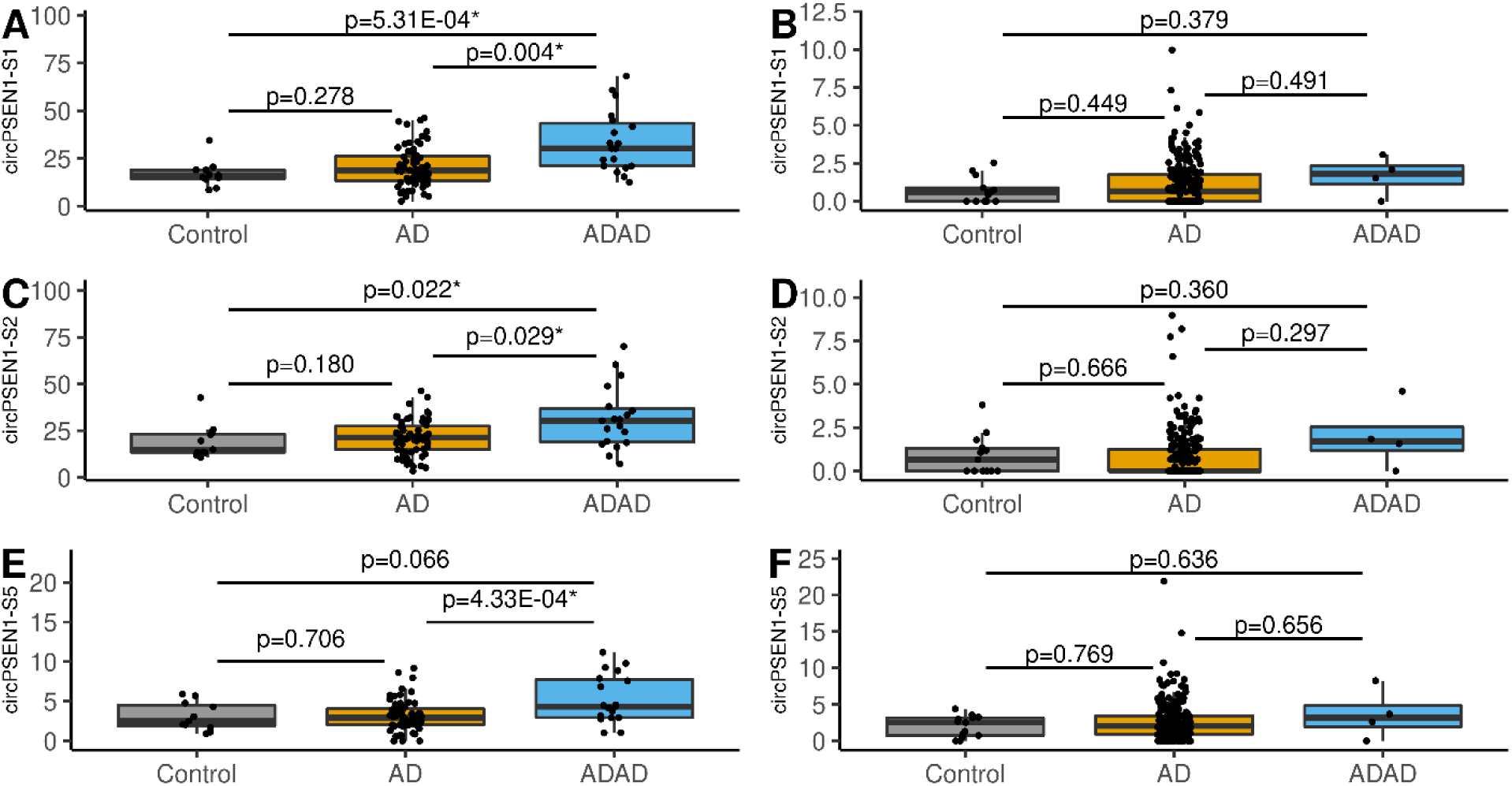
Comparison of the circular *PSEN1* normalized counts for the three main *circPSEN1* species S1 - hsa_circ_0008521 (Panel A in discovery, Panel B in replication), S2-hsa_circ_0003848 (Panel C in discovery, Panel D in replication), and S5 - hsa_circ_0002564 (Panel E in discovery, Panel F in replication) for Controls (grey), AD (Alzheimer disease - ocher) and ADAD (autosomal dominant Alzheimer disease - blue)

**Supplementary Figure 3.**
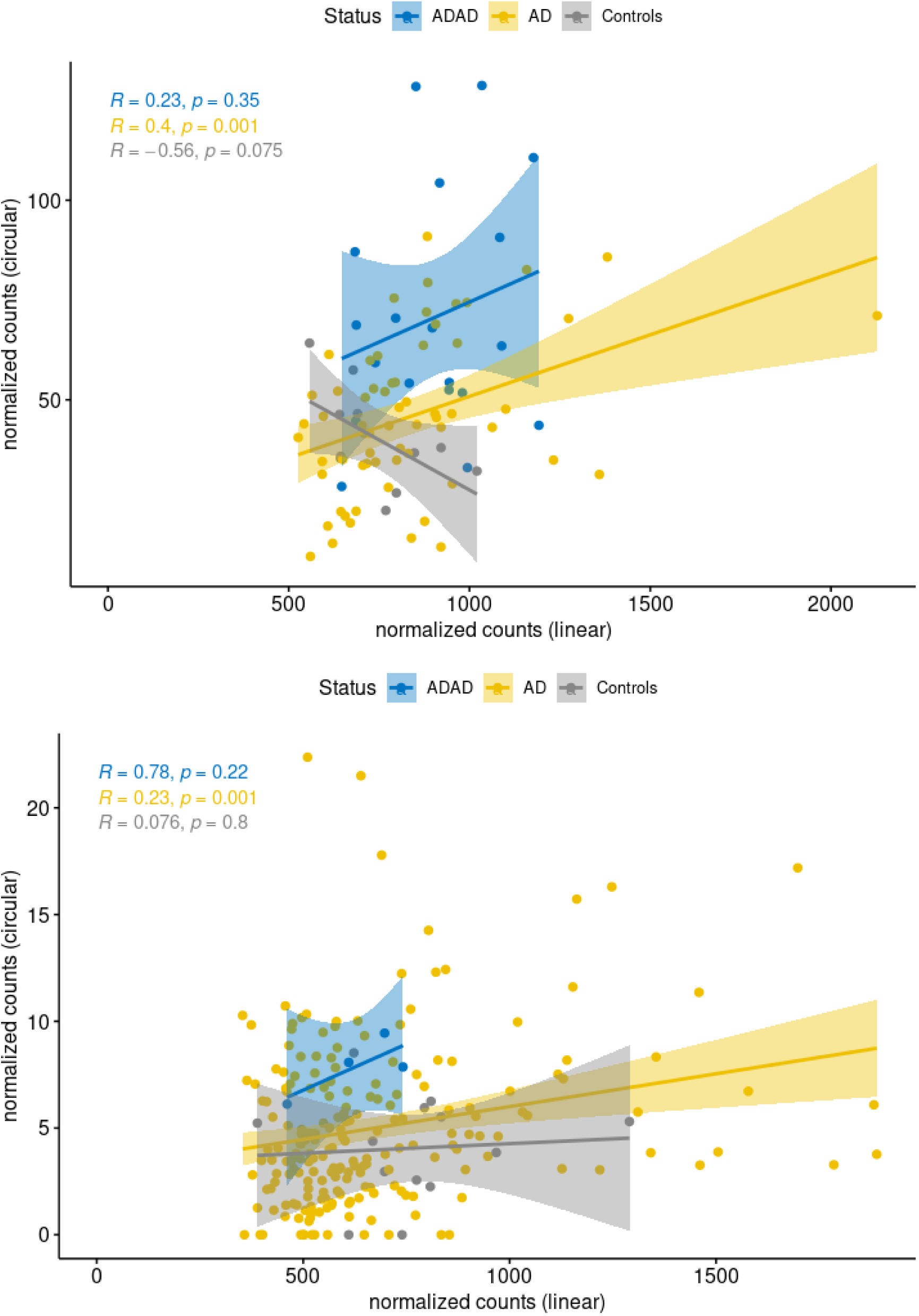
Correlation of circular *PSEN1* normalized counts and linear *PSEN1* normalized counts in ADAD (blue), AD (yellow), and controls (gray) for the discovery (Panel A) and replication (Panel B) Datasets

**Supplementary Figure 4.**
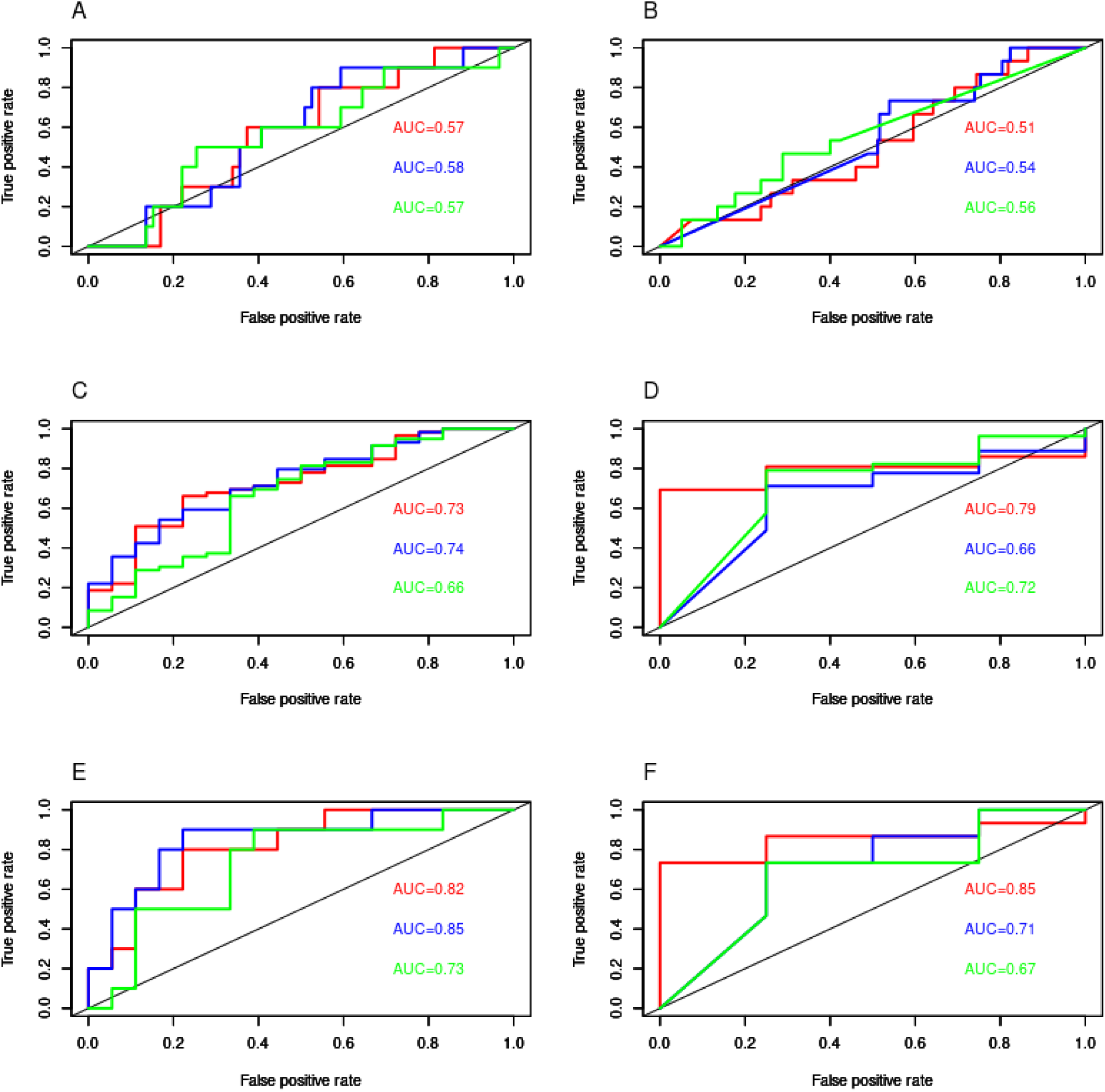
ROCs corresponding to the binomial regression models using different circ*PSEN1* counts (aggregate - red, hsa_circ_0008521 - blue, hsa_circ_0003848 - green) for classifying AD vs Controls (Panel A in discovery and Panel B in replication), ADAD vs AD (Panel C in discovery and Panel D in replication), ADAD vs Controls (Panel E in discovery and Panel F in replication).

